# Repurposing probucol for prevention of dementia: Evidence from a nationwide cohort study

**DOI:** 10.1101/2025.11.17.25340427

**Authors:** Ryuske Takechi, Jennifer Dunne, Virginie Lam, Blossom Christa Maree Stephan, Gavin Pereira, Roger Clarnette, Gerald Francis Watts, Leon Flicker, Suzanne Robinson, Christopher Michael Reid, Arazu Sharif, Sean Randall, John Charles Louis Mamo

## Abstract

**BACKGROUND:** Effective treatments for neurodegenerative diseases remain elusive, underscoring the importance of preventive strategies. Probucol, a cholesterol lowering and antioxidant drug with established cardiovascular use, has shown neuroprotective effects in preclinical models of dementia by modulating peripheral lipoprotein amyloid metabolism and preserving capillary integrity. However, no large-scale human studies have examined its association with dementia risk.

**OBJECTIVE:** To examine the association between probucol use and incident dementia in older adults.

**DESIGN, SETTING, AND PARTICIPANTS:** This retrospective cohort study used the Japan Medical Data Centre claims database from 2014 to 2023. Adults aged 50 years or older prescribed probucol or statins were included, excluding those with prior dementia or recent drug exposure. Participants were categorized as probucol monotherapy users, statin monotherapy users, or combination users (≥2 prescriptions). Propensity score matching was used to balance baseline comorbidities.

**EXPOSURES:** Probucol or statin therapy.

**MAIN OUTCOMES AND MEASURES:** The primary outcome was incident all cause dementia. Secondary outcomes included Alzheimer’s disease and mixed Alzheimer’s and vascular dementia. Odds ratios (ORs) with 95% CIs were calculated using logistic regression adjusted for age and sex.

**RESULTS:** Among 57 231 individuals (52.6% female) followed for up to 10 years (median, 3 years), 7 387 (12.9%) developed dementia. The cohort included 2 896 probucol users (5.1%) and 54 335 statin users (94.9%). Dementia incidence was higher among females (14.7%) than males (10.9%). Dementia incidence was lower in probucol users (5.6%, 162/2 896) than in statin users overall (13.2%, 8 248/62 519), with individual statins ranging from 11.4% (fluvastatin) to 16.7% (pravastatin). Probucol use was associated with a 62% lower adjusted risk of dementia compared with all statin users combined (adjusted OR, 0.38; 95% CI, 0.37-0.38). Protective associations were consistent across individual statin comparisons, with adjusted ORs ranging from 0.30 (pitavastatin) to 0.57 (fluvastatin).

**CONCLUSIONS AND RELEVANCE:** In this large national cohort of Japanese adults, probucol use was associated with a substantially lower risk of incident dementia compared with statins. These findings provide the first large-scale human evidence linking probucol exposure with reduced dementia risk, supporting its further evaluation as a preventive therapy in prospective clinical trials.

**Key Points:** *Question:* Is use of the lipid-lowering and antioxidant agent probucol associated with a reduced risk of incident dementia compared with statin therapy in older adults?

*Findings:* In this nationwide cohort study of 57 231 Japanese adults aged 50 years and older, dementia incidence was nearly halved in probucol users (5.6%) compared with statin users overall (13.2%), corresponding to 62% lower adjusted odds of dementia. Protective associations were consistent across dementia subtypes and individual statins.

*Meaning:* These findings suggest that probucol, a long-standing and well-tolerated cardiovascular drug, may offer a mechanistically distinct and potentially scalable strategy for dementia prevention, warranting confirmation in prospective intervention trials.

## Introduction

Dementia and mild cognitive impairment represent a growing global health crisis, with more than 57 million people affected worldwide and nearly 10 million new cases each year.^1^ Despite substantial advances in understanding disease mechanisms, effective disease-modifying treatments remain elusive, underscoring the urgent need for preventive strategies.^2^ Traditionally, dementia research has focused on brain-derived amyloid and tau pathology; however, mounting evidence implicates systemic vascular and metabolic disturbances as upstream contributors to neurodegeneration.^3–7^

More than 90% of circulating amyloid-β (Aβ) is bound to lipoproteins, particularly triglyceride-rich lipoproteins (TRLs) of intestinal and hepatic origin.^3,8^ Dysregulated metabolism of these amyloid-associated lipoproteins has been linked to endothelial injury, blood–brain barrier disruption, neurovascular inflammation, and cognitive decline in both preclinical and human studies.^4–6,9,10^ This emerging “lipoprotein–amyloid axis” offers a plausible mechanistic bridge between systemic lipid metabolism and neurodegenerative processes.

Probucol, a lipid-lowering agent still prescribed in parts of Asia for secondary cardiovascular prevention, exhibits pleiotropic actions of potential relevance to dementia beyond cholesterol reduction.^11^ Compared with statins, probucol profoundly suppresses TRL-associated Aβ biosynthesis and secretion, preserves blood–brain barrier integrity and attenuates neurovascular inflammation.^9,12–14^ It is also one of the most potent small-molecule antioxidants in clinical use.^15^ These convergent actions suggest that probucol may modify disease processes at the neurovascular interface, a property not shared by more potent contemporary lipid-lowering therapies..

In this nationwide claims-based cohort study in Japan, we evaluated dementia risk in patients with cardiovascular disease treated with probucol (with or without statins) compared with those treated with statins alone. We hypothesised that probucol use would be associated with a lower risk of incident dementia. These findings provide the first large-scale human evidence supporting a peripheral, lipoprotein-mediated contribution to dementia pathogenesis and identify probucol as a candidate for repurposing in dementia prevention.

## Methods

### Data Source

We conducted a retrospective cohort study using the Japan Medical Data Centre (JMDC) claims database,^16^ which contains comprehensive health data from employed individuals and their dependents enrolled in health insurance societies. The database includes anonymised information on demographics, diagnoses (using International Classification of Diseases, 10th Revision [ICD-10] codes), prescriptions, procedures and healthcare utilisation. This study contains data from 2014 to 2023 and over 27 million individuals.

### Study Population

We identified individuals aged ≥50 years who had been prescribed probucol or a statin drug (atorvastatin; fluvastatin; pitavastin; pravastatin; rosuvastatin; simvastatin). Individuals were followed from January 1, 2014, to December 31, 2023.

### Exposure Definition

Drug exposure was defined based on prescription records, requiring a minimum of two filled prescriptions to ensure sustained use and minimise misclassification bias. We categorized participants into three mutually exclusive exposure groups: probucol monotherapy users (≥2 probucol prescriptions without concurrent statin use), combination users (≥2 prescriptions for both probucol and statins) and statin monotherapy users (≥2 statin prescriptions without probucol use). This classification approach captured both single-agent and combination therapy patterns while accounting for the clinical reality that patients may switch between or combine lipid-lowering therapies over time. For secondary dose-response analyses, probucol exposure was further stratified by treatment duration using a 120-day cut-off to define short-term (<120 days) versus long-term (≥120 days) exposure.

### Outcome Assessment

The primary outcome was incident all-cause dementia, defined using comprehensive diagnostic criteria based on standardized disease names in the JMDC database rather than traditional ICD-10 codes. Our dementia definition encompassed a broad spectrum of dementia-related conditions including Alzheimer’s disease (familial, presenile, senile and atypical variants), vascular dementia (including multi-infarct and acute onset forms), mixed dementia, dementia with Lewy bodies, frontotemporal dementia, Parkinson’s disease with dementia, cortical and subcortical dementia, alcoholic dementia, HIV dementia and other secondary dementia types. The definition also captured related neurodegenerative conditions such as Huntington disease dementia, CADASIL, CARASIL, argyrophilic grain dementia and senile brain degeneration.

Secondary outcomes included specific dementia subtypes for subgroup analyses: (1) Alzheimer’s disease, defined as cases with Alzheimer’s disease diagnoses (including familial, presenile, senile and atypical variants) without concurrent vascular dementia diagnoses; and (2) mixed Alzheimer’s disease with vascular dementia, defined as cases with diagnoses of both Alzheimer’s disease and vascular dementia components.

### Covariates

We extracted baseline covariates across four domains to characterize study participants and adjust for potential confounding. Demographic variables included age stratified into four categories (50-59, 60-69, 70-79 and ≥80 years) and binary gender classification (male, female). Comorbidity status was determined using comprehensive ICD-10 diagnostic codes to identify five key conditions: hypertension (including essential, pulmonary, renovascular and secondary forms), diabetes mellitus (encompassing type 1, type 2, steroid-induced and pancreatic variants), lipid disorders (hyperlipidemia, hypercholesterolemia, dyslipidemia and familial conditions), angina (angina pectoris and arteriosclerosis obliterans) and hepatic disorders (hepatic dysfunction, steatosis, fibrosis and related hepatic conditions).

### Propensity Score Matching

To address potential confounding by indication, we employed propensity score matching to balance baseline characteristics between exposure groups.^17^ Propensity score matching was performed at the individual level using aggregated baseline data derived from record-level claims. We conducted PSM with nearest neighbour matching, with all baseline covariates achieved standardised mean differences <0.1 after matching, indicating excellent balance between groups.^18^ The propensity score model included key comorbidities (hypertension, diabetes, lipid disorders, angina and hepatic disorders) as matching variables. We matched each probucol user to four non-users with the nearest propensity scores using a calliper width of 0.2 of the standard deviation of the logit of the propensity score, except for simvastatin which was matched 1:2 due to low drug user numbers.

Our starting cohort population was 77,312 individuals (Figure 1). After excluding individuals younger than 50 years (n=6,477, 8.4%), individuals who were prescribed drugs within 12 months of baseline (n=13,382, 17.3%) and individuals who had a dementia-related records within 12 months of base (n=126, <1%). After these exclusions, the total number of eligible individuals for inclusion in the study is 57,231 (Figure 1). All subsequent statistical analyses were performed on this propensity score-matched cohort.

**Figure 1.**
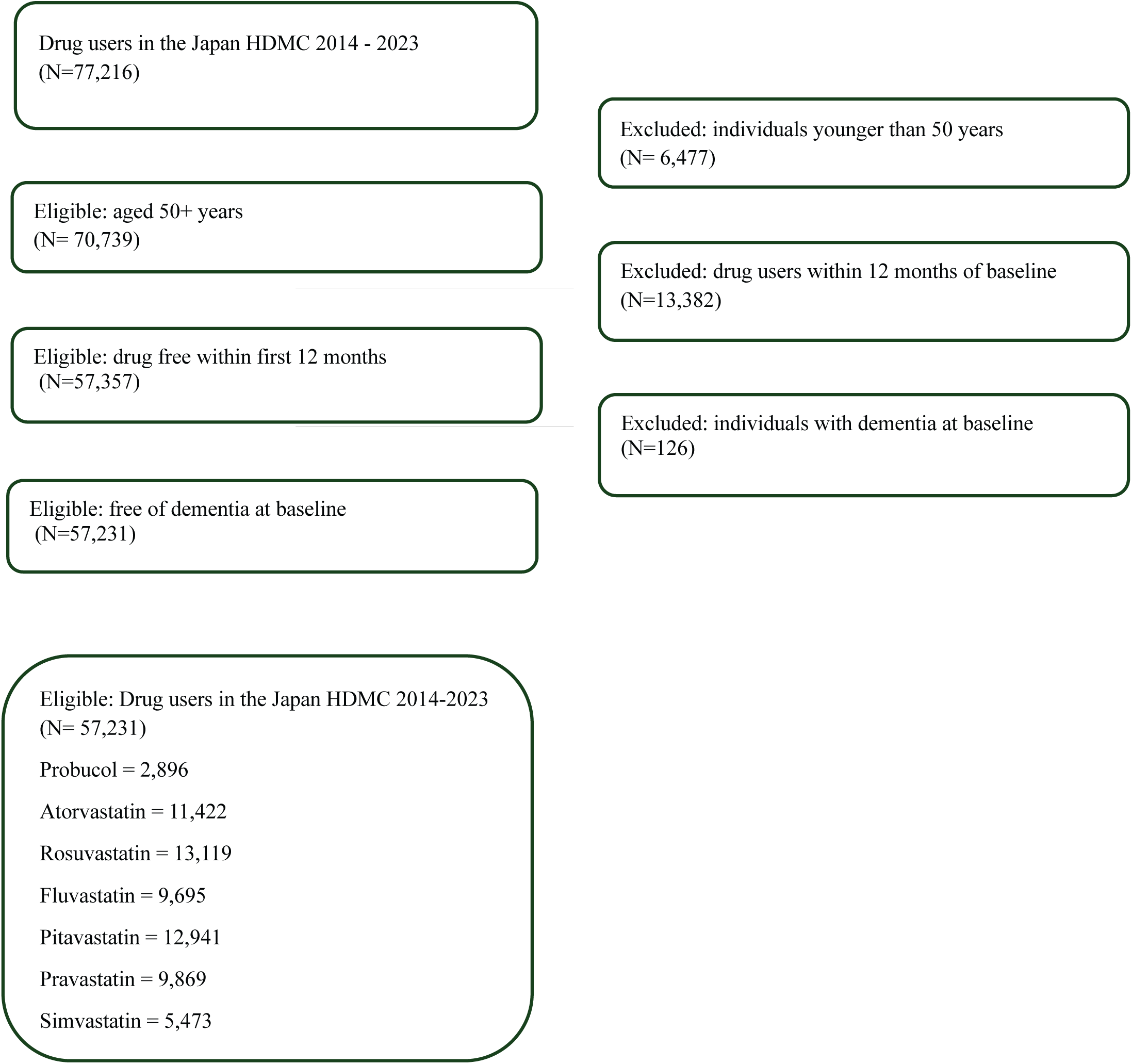
Selection of eligible health claims records included in this study, Japan HDMC 2014-2023.

### Statistical Models

We conducted logistic regression analyses on the propensity score-matched cohort to examine the association between probucol use and incident dementia risk for both the primary outcome (all-cause dementia) and secondary outcomes (Alzheimer’s disease subtype and mixed Alzheimer’s disease with vascular dementia subtype). Drug exposure was defined using prescription records, with participants categorized into mutually exclusive groups: probucol monotherapy users, individual statin monotherapy users (atorvastatin, rosuvastatin, fluvastatin, pitavastatin, pravastatin, simvastatin) and combination users.

We implemented two analytical approaches: first, comparing probucol users against all statin users combined as a single reference group; second, conducting individual comparisons between probucol users and each specific statin type separately. In both approaches, the 1:4 propensity score matching structure was maintained, with each probucol user retaining their four matched statin controls from the primary matching procedure. For each comparison, we fitted both unadjusted and adjusted logistic regression models. Adjusted models included age (continuous), sex (male/female), while unadjusted models examined the crude association between drug exposure and dementia risk. We excluded patients using multiple statins simultaneously to maintain clear exposure classifications.

For subgroup analyses, we repeated the same analytical approach separately for participants who developed Alzheimer’s disease and for those who developed mixed Alzheimer’s disease with vascular dementia, using the same exposure definitions and adjustment variables. To ensure sustained drug exposure and minimise misclassification bias, we excluded patients with treatment gaps ≥120 days between consecutive prescriptions, a threshold commonly used in pharmacoepidemiological studies to distinguish between temporary treatment interruptions and true discontinuation.^19^

Odds ratios (ORs) with 95% confidence intervals were calculated to quantify the association between probucol use and dementia risk. For the individual statin comparisons, we systematically releveled each statin as the reference category to generate comprehensive pairwise comparisons with probucol. All analyses were performed using logistic regression generalised linear model with binomial family and logit link and results were exponentiated to obtain interpretable odds ratios. The analysis focused on patients aged ≥50 years with complete demographic and prescription data, ensuring robust estimation of treatment effects across different lipid-lowering therapeutic approaches.

All analyses were performed using R version 4.4.3 (R Foundation for Statistical Computing, Vienna, Austria).

## Results

### Baseline Characteristics

Of the 57,231 individuals aged ≥50 years included in the analysis 52.6% were female (n=30,111) and 47.4% male participants (n=27,120) (Table 1). The median age distribution showed the largest representation in the 70-74 years age group, with 5,247 females and 5,319 males, representing 18.5% of the total cohort. Age distribution was relatively balanced across sexes, with participants ranging from 50 to >100 years.

**Table 1.** Characteristics of the study population (n= 57,231) in the Japan HDMC who were prescribed probucol and statin drugs between 2014 and 2023.

| **Characteristics** | **Total (%)**<br>57,231 | **Female N (%)**<br>30,111 | **Male N (%)**<br>27,120 |
| --- | --- | --- | --- |
| **Age (years):** |  |  |  |
| 50-54 | 3,234 (5.7%) | 1,461 (4.9%) | 1,773 (6.5%) |
| 55-59 | 4,435 (7.8%) | 2,009 (6.7%) | 2,426 (9.0%) |
| 60-64 | 6,874 (12.0%) | 3,193 (10.6%) | 3,681 (13.6%) |
| 65-69 | 9,754 (17.0%) | 4,714 (15.7%) | 5,040 (18.6%) |
| 70-74 | 10,566 (18.5%) | 5,247 (17.4%) | 5,319 (19.6%) |
| 75-79 | 9,771 (17.1%) | 5,383 (17.9%) | 4,388 (16.2%) |
| 80-84 | 7,530 (13.2%) | 4,650 (15.4%) | 2,880 (10.6%) |
| 85-89 | 3,810 (6.7%) | 2,491 (8.3%) | 1,319 (4.9%) |
| 90-94 | 1,054 (1.8%) | 788 (2.6%) | 266 (<1%) |
| 95-100 | 90 (<1%) | 76 (<1%) | 14 (<1%) |
| >100 | 113 (<1%) | 99 (<1%) | 14 (<1%) |
| **Drug:** |  |  |  |
| Probucol | 2,896 (5.1%) | 1,588 (5.3%) | 1,308 (4.8%) |
| Atorvastatin | 11,422 (20.0%) | 5,792 (19.2%) | 5,630 (20.8%) |
| Fluvastatin | 9,695 (17.0%) | 5,449 (18.1%) | 4,246 (15.7%) |
| Pitavastatin | 12,941 (22.6%) | 6,245 (20.7%) | 6,696 (24.7%) |
| Pravastatin | 9,869 (17.2%) | 5,775 (19.2%) | 4,094 (15.1%) |
| Rosuvastatin | 13,119 (22.9%) | 5,930 (19.7%) | 7,189 (26.5%) |
| Simvastatin | 5,473 (9.7%) | 3,504 (11.6%) | 1,969 (7.3%) |
| **Outcome cases:** |  |  |  |
| All cause dementia | 7,387 (12.9%) | 4,439 (14.7%) | 2,948 (10.9%) |
| Alzheimer's disease | 3,715 (6.5) | 2,348 (7.8%) | 1,367 (5.0%) |
| Vascular dementia | 198 (<1%) | 90 (<1%) | 108 (<1%) |

Among the study population, 2,896 individuals (5.1%) were prescribed probucol, while the remaining participants received various statin medications. Statin prescriptions were distributed as follows: rosuvastatin (n=13,119, 22.9%), pitavastatin (n=12,941, 22.6%), atorvastatin (n=11,422, 20.0%), pravastatin (n=9,869, 17.2%), fluvastatin (n=9,695, 16.9%) and simvastatin (n=5,473, 9.6%).

During a median follow-up of 3 years (maximum 10 years), 7,387 individuals (12.9%) developed incident all-cause dementia, with a higher proportion among females (n=4,439, 14.7%) compared to males (n=2,948, 10.9%). Within this group, 3,715 cases were Alzheimer’s disease (6.5% of the cohort) and 198 were vascular dementia (0.3%). Dementia incidence was lower in probucol users (5.6%, 162/2,896) compared to statin users overall (13.2%, 8,248/62,519), with individual statins ranging from 11.4% (fluvastatin) to 16.7% (pravastatin) (Figure 2).

**Figure 2.**
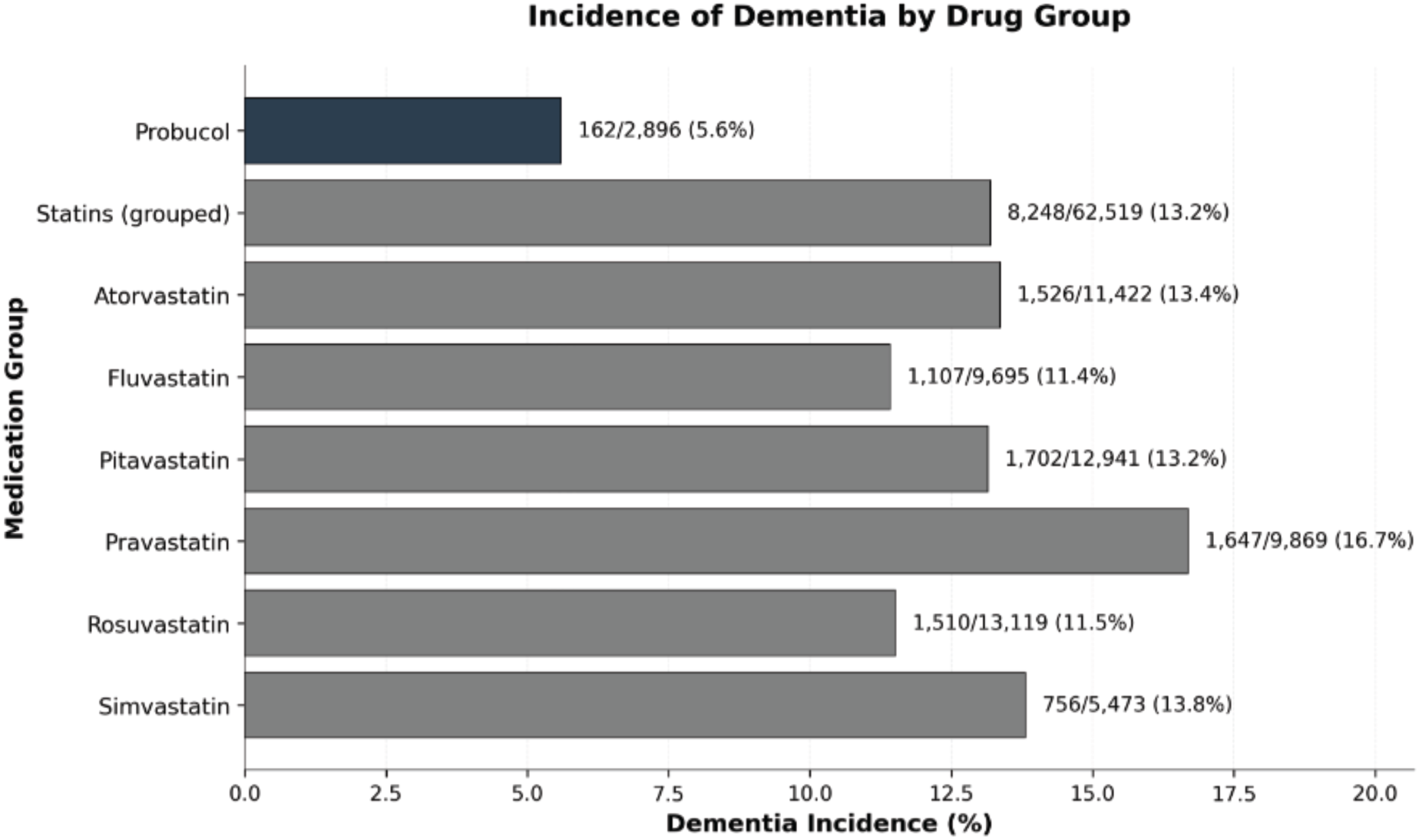
Incidence of dementia among patients receiving probucol versus statin therapy in the Japan HDMC (n= 57,231)

#### A ssociation Between Probucol Use and Dementia Risk

When comparing probucol users to all statin users combined, probucol was associated with a significantly lower risk of all-cause dementia (Table 2). The unadjusted odds ratio was 0.43 (95% CI: 0.43-0.44), which remained significant after adjustment for age and sex (adjusted OR [aOR] 0.38, 95% CI 0.37–0.38), corresponding to a 62% risk reduction.

**Table 2.** Unadjusted and adjusted odds ratio for the association between probucol and statin drugs and the development of dementia in the Japan HDMC (n= 57,231)

| Drug | Dementia |  | Alzheimer disease |  | Alzheimer disease with vascular dementia |  | Dementias not including Alzheimer disease |  |
| --- | --- | --- | --- | --- | --- | --- | --- | --- |
|  | Unadjusted OR (95% CI) | Adjusted OR (95% CI) | Unadjusted OR (95% CI) | Unadjusted OR (95% CI) | Adjusted OR (95% CI) | Adjusted OR (95% CI) | Unadjusted OR (95% CI) | Adjusted OR (95% CI) |
| **Probucol** | 0.43 (0.35 – 0.53) | 0.37 (0.29 – 0.45) | 0.50 (0.38 – 0.65) | 0.49 (0.37 – 0.64) | 0.44 (0.33 – 0.57) | 0.44 (0.33 – 0.58) | 0.41 (0.31 – 0.52) | 0.36 (0.28 – 0.47) |
| **Statin** | Reference | Reference |  |  |  |  |  |  |
| **Probucol** | 0.43 (0.34-0.52) | 0.33 (0.26 – 0.41) | 0.59 (0.44 – 0.78) | 0.58 (0.43 – 0.76) | 0.48 (0.35 – 0.63) | 0.49 (0.36 – 0.65) | 0.36 (0.27 – 0.47) | 0.29 (0.22 – 0.38) |
| **Atorvastatin** | Reference | Reference |  |  |  |  |  |  |
| **Probucol** | 0.53 (0.42 – 0.65) | 0.57 (0.47 – 0.71) | 0.53 (0.39 – 0.69) | 0.52 (0.39 – 0.68) | 0.57 (0.42 – 0.76) | 0.58 (0.43 – 0.77) | 0.58 (0.43 – 0.75) | 0.63 (0.47 – 0.83) |
| **Fluvastatin** | Reference | Reference |  |  |  |  |  |  |
| **Probucol** | 0.44 (0.35 – 0.54) | 0.30 (0.24 – 0.37) | 0.52 (0.39 – 0.69) | 0.52 (0.39 – 0.68) | 0.37 (0.28 – 0.49) | 0.37 (0.28 – 0.49) | 0.52 (0.39 – 0.68) | 0.37 (0.28 – 0.49) |
| **Pitavastatin** | Reference | Reference |  |  |  |  |  |  |
| **Probucol** | 0.31 (0.25 – 0.38) | 0.31 (0.25 – 0.39) | 0.37 (0.28 – 0.49) | 0.37 (0.27 – 0.48) | 0.39 (0.29 – 0.51) | 0.39 (0.29 – 0.52) | 0.30 (0.22 – 0.38) | 0.31 (0.23 – 0.40) |
| **Pravastatin** | Reference | Reference |  |  |  |  |  |  |
| **Probucol** | 0.51 (0.41 – 0.63) | 0.32 (0.25 – 0.37) | 0.63 (0.47 – 0.83) | 0.62 (0.46 – 0.81) | 0.40 (0.29 – 0.53) | 0.40 (0.30 – 0.54) | 0.45 (0.34 – 0.58) | 0.30 (0.22 – 0.39) |
| **Rosuvastatin** | Reference | Reference |  |  |  |  |  |  |
| **Probucol** | 0.41 (0.33 – 0.51) | 0.51 (0.40 – 0.64) | 0.41 (0.30 – 0.54) | 0.41 (0.30 – 0.54) | 0.50 (0.37 – 0.67) | 0.51 (0.37 – 0.68) | 0.46 (0.35 – 0.61) | 0.56 (0.42 – 0.74) |
| **Simvastatin** | Reference | Reference |  |  |  |  |  |  |

\*Adjusted for age (continuous) and sex

In head-to-head comparisons with individual statins, probucol consistently demonstrated protective associations, though effect sizes varied (Figure 3). The strongest protective association was observed compared to pravastatin users (aOR = 0.31, 95% CI: 0.24-0.38), representing a 69% risk reduction, followed by pitavastatin (aOR = 0.30, 95% CI: 0.24-0.38) with a 70% risk reduction. Substantial protective effects were also observed compared to atorvastatin (aOR = 0.33, 95% CI: 0.26-0.41) and rosuvastatin (aOR = 0.34, 95% CI: 0.27-0.42), both showing approximately 67-66% risk reductions. More modest but still significant protective effects were observed compared to simvastatin (aOR = 0.51, 95% CI: 0.40-0.64) with a 49% risk reduction and fluvastatin (aOR = 0.57, 95% CI: 0.45-0.71) with a 43% risk reduction.

**Figure 3.**
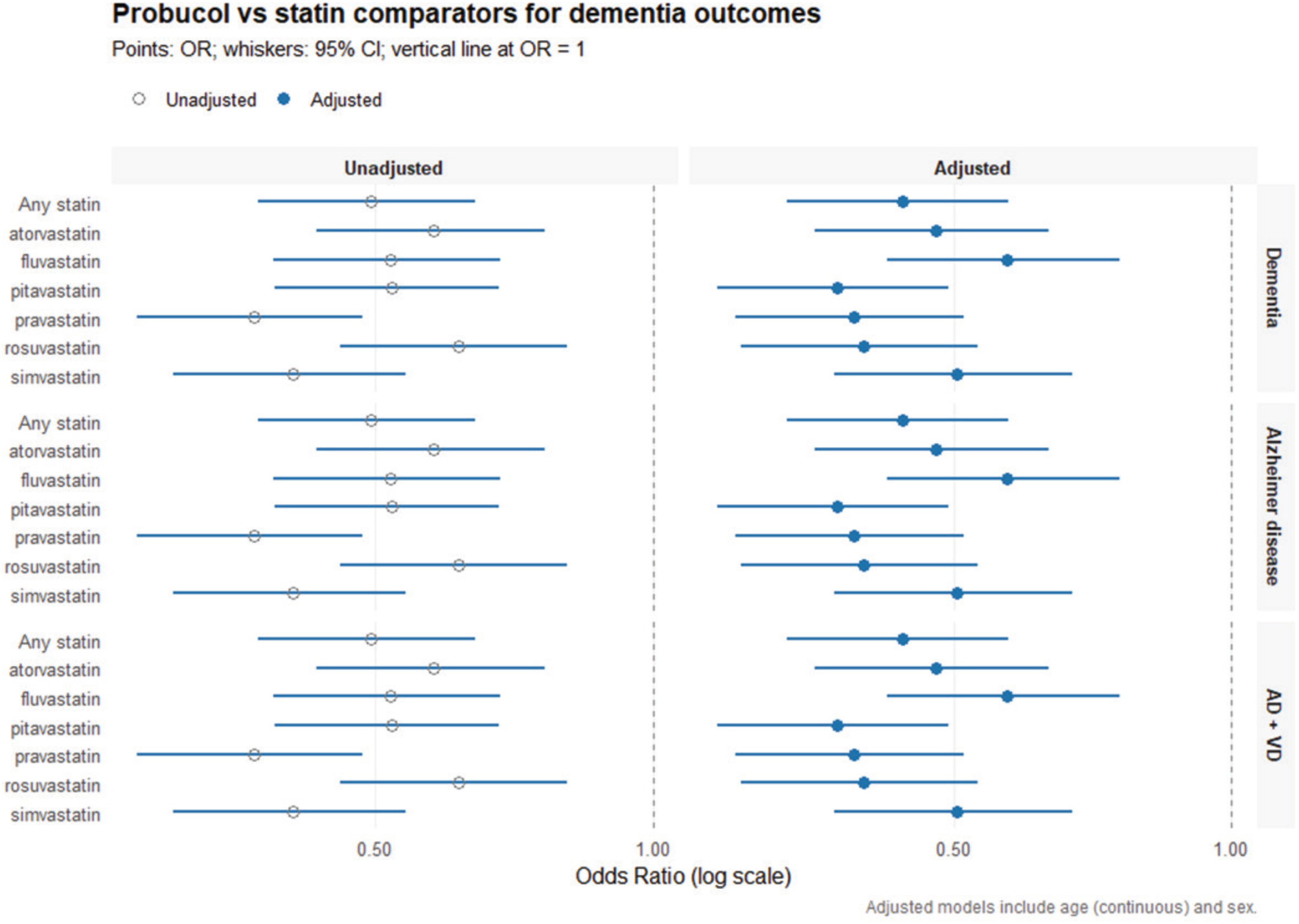
Unadjusted and adjusted odds ratio for the association between probucol and statin drugs and the development of dementia in the Japan HDMC (n= 57,231)

For Alzheimer’s disease specifically, probucol showed consistent protection across all comparisons, with adjusted ORs ranging from 0.37 (95% CI 0.28–0.49; vs. pitavastatin) to 0.58 (95% CI 0.43–0.77; vs. fluvastatin). Similar patterns were observed for mixed Alzheimer’s and vascular dementia, with adjusted ORs ranging from 0.37 (95% CI 0.28–0.49) versus pitavastatin to 0.57 (95% CI 0.42–0.76) versus fluvastatin.

### Sensitivity Analysis

Sensitivity analysis excluding participants with probucol inter-prescription intervals exceeding 120 days (n=414) showed concordant results with the primary analysis (Supplementary Table 1). Probucol remained highly protective against dementia across all comparisons, with adjusted ORs ranging from 0.30 (vs. pitavastatin) to 0.57 (vs. fluvastatin), confirming the robustness of findings irrespective of treatment continuity.

## Discussion

This large-scale retrospective cohort study provides the first human evidence that probucol, an agent with polypharmacological properties, is associated with a substantially reduced risk of dementia compared with statin therapy alone. Our findings demonstrate a consistent 62% reduction in all-cause dementia risk among probucol users, with protective effects observed across individual statin comparisons and dementia subtypes, including Alzheimer’s disease and mixed vascular–Alzheimer’s dementia. These results lend strong support to the emerging paradigm that peripheral lipoprotein–amyloid metabolism plays a central role in dementia pathogenesis, providing a plausible mechanistic rationale for probucol’s neuroprotective effects that extends beyond traditional cholesterol-lowering pathways. The magnitude and consistency of these associations, observed across a diverse population of more than 57,000 individuals followed for up to 10 years, suggest that probucol’s unique biological actions may translate into clinically meaningful cognitive protection in real-world settings.

Preclinical studies have consistently shown that exaggerated lipoprotein–amyloid metabolism contributes to blood–brain barrier (BBB) breakdown, extravasation of amyloid-laden lipoproteins into the parenchyma, neurovascular inflammation and neurodegeneration that precedes plaque formation.^5–7,20^ Probucol appears to act at multiple points along this pathogenic cascade.^21^ It not only suppresses the secretion of amyloid with nascent lipoproteins but also accelerates its peripheral clearance, thereby reducing capillary exposure. Probucol has also been shown to stimulate endothelial tight junction protein expression, strengthening the BBB and limiting parenchymal insult from circulating proteins and macromolecules.^13,14,22^ In addition, probucol beneficially modulates key inflammatory enzymes including heme oxygenases, superoxide dismutase and glutathione peroxidase, while exerting potent direct antioxidant activity.^21,23^ Together, these actions converge at the neurovascular interface to mitigate the systemic–central pathology that underpins dementia progression. The particularly strong associations observed for non-Alzheimer’s dementia subtypes in this study may reflect the importance of vascular and metabolic pathways in their aetiology, where probucol’s multimodal effects are especially relevant.

Epidemiological studies and meta-analyses evaluating statins for dementia prevention have yielded mixed results.^24^ Although several reports suggest a modest protective association with Alzheimer’s disease and all-cause dementia, typically reflecting an odds ratio reduction of about 0.2, other studies have not demonstrated significant benefit. The modest and variable effect observed with statins contrasts with the magnitude of risk reduction identified here for probucol. Mechanistically, statins primarily lower LDL-cholesterol and exert limited influence on the metabolism of amyloid-enriched lipoproteins such as very low-density lipoproteins (VLDL) and chylomicrons.^25^ In contrast, probucol’s broader metabolic actions, including suppression of amyloid secretion with nascent triglyceride-rich lipoproteins, preservation of blood–brain barrier integrity and attenuation of neurovascular inflammation, may underlie its stronger association with reduced dementia risk.^14,21,22^ These findings extend existing literature by identifying a distinct therapeutic avenue that warrants evaluation in prospective interventional studies.

An important consideration is that our study population comprised individuals with established cardiovascular disease, a group already at several-fold higher risk of dementia owing to underlying vascular pathology. This makes the consistent and substantial risk reduction observed particularly noteworthy, as it suggests that probucol can confer protection even in a population that is comparatively harder to treat. By extension, probucol may be even more effective in individuals with a lower vascular burden, in whom neurovascular injury is less advanced and protective mechanisms may be more readily preserved. Given that most dementia cases arise in the general population without overt cardiovascular disease, these findings raise the possibility that probucol could offer even greater preventive benefit when introduced earlier in the disease trajectory, before cumulative vascular injury and neurodegeneration take hold.

If validated in prospective trials such as the *Probucol in Alzheimer’s Study* (ACTRN12621000726853),^11^ these findings have important clinical implications. Probucol is an inexpensive, orally available agent with a long history of safe use across several Asian nations including Japan, South Korea, Taiwan, China, India and parts of Southeast Asia such as Thailand and the Philippines, where it remains licensed for secondary cardiovascular prevention. Its multimodal actions at the vascular–amyloid interface make it a promising candidate for repurposing as a dementia-preventive therapy. The effect sizes observed in this study suggest that probucol could confer meaningful risk reduction, particularly in populations with an elevated vascular burden. From a public health perspective, targeting systemic lipoprotein–amyloid metabolism represents a novel and potentially scalable approach to dementia prevention that complements existing strategies focused on brain-derived amyloid and tau pathology.

### Limitations

This study has several notable strengths. The large sample size of over 57,000 individuals from a comprehensive national claims database with up to 10 years of observation and a median of 3 years of follow-up allows for robust assessment of dementia outcomes. Our propensity score–matching approach effectively balanced baseline observed characteristics between treatment groups, addressing confounding by indication that commonly affects observational pharmacoepidemiology. The use of person-level aggregated data, rather than record-level analysis, ensured methodological validity by avoiding bias from non-independent observations and exposure misclassification. Additionally, our comprehensive dementia definition, based on standardized disease names, captured a broad spectrum of dementia subtypes, increasing clinical relevance beyond the limitations of conventional ICD-10 coding.

Several caveats merit consideration. Despite the overall sample size, the relatively small number of probucol users (n = 2,896, 5.1%) may have limited statistical power for detecting modest effect sizes in subgroup analyses. The observational design cannot establish causality and residual confounding from unmeasured factors remains possible. As with claims database studies, our analysis was limited to variables captured for administrative and billing purposes, which excluded key potential confounders such as socioeconomic variables (education, income, occupation), lifestyle factors (smoking, alcohol use, physical activity, diet), body mass index, and cognitive test scores. Our exposure definition required at least two prescriptions, which may have excluded individuals with very brief trials of probucol, though this approach was necessary to ensure sustained exposure. The relatively short median follow-up of 3 years may not capture the full trajectory of dementia risk, particularly given the lengthy prodromal phase of neurodegenerative disease. We also lacked data on treatment adherence beyond prescription fills and on clinical severity markers that may have influenced prescribing decisions. Although propensity matching addressed measured confounders, unmeasured confounding by indication cannot be fully excluded, particularly given the specific clinical contexts in which probucol is prescribed in Japan. Finally, generalisability to non-Japanese populations remains uncertain, as differences in genetic background, lifestyle and healthcare systems could modify treatment effects.

## Conclusion

This nationwide cohort analysis provides compelling human evidence that probucol use is associated with a substantially reduced risk of dementia, reinforcing a mechanistic link between systemic metabolic regulation and neurovascular protection. While causal inference cannot be established from observational data, the magnitude and consistency of the associations observed together with convergent preclinical evidence justify formal evaluation of probucol in randomized prevention trials. The ongoing *Probucol in Alzheimer’s Study (PIA-Study)* (ACTRN12621000726853) and the development of next-generation probucol analogues represent important next steps toward defining the therapeutic potential of this class for dementia prevention and vascular brain health.

## Data Availability

The data that support the findings of this study were obtained from the Japan Medical Data Centre (JMDC) under a formal data-use agreement and cannot be shared publicly due to licensing restrictions. Aggregate data and analytic code are available from the corresponding author upon reasonable request, subject to JMDC approval and institutional ethics requirements.

## Acknowledgements

The authors would like to thank the Bryant Stokes Neurological Research Fund (BSNRF), the McCusker Charitable Foundation, Wen Giving Foundation and MSWA for funding the project and the data custodians of the Japan Medical Data Centre (JMDC) claims database for providing the health data for this study.

## Contributors

JM and RT conceived and designed the study. JD undertook the data curation, data analysis and drafted the initial version of the manuscript with JM and RT. All authors contributed to the interpretation of the data and critically revised the manuscript. All authors had full access to the tables and figures in the study and can take responsibility for the integrity of the data and the accuracy of the data analysis. JM, the corresponding author, attests that all listed authors meet authorship criteria and that no other meeting the criteria were excluded.

## Ethical approval

This study was conducted in accordance with the principles of the Declaration of Helsinki and followed company-specific guidelines for database analysis. Given the anonymized nature of the data, the requirement for written informed consent and ethical approval for this study were waived by the Human Research Ethics Committee, Curtin University (EXTL2024-003).

## Data sharing

The Japan Medical Data Centre (JMDC) claims database owned by the data custodians for the Japan Medical Data Centre who approved the use of the data for this study. Use of the study data is restricted to named researchers. The current Human Research Ethics Committee approvals were obtained for public sharing and presentation of data on group level only, meaning the data used in this study cannot be shared by the authors. The steps involved in seeking permission for use of the data used in this study are the same for all researchers. The lead authors (JM, RT and JD) affirms that the manuscript is an honest, accurate, and transparent account of the study being reported. No important aspects of the study have been omitted, and any discrepancies from the study as originally planned have been explained.

